# Cerebrospinal fluid p-tau217 performs better than p-tau181 as a biomarker of Alzheimer’s disease

**DOI:** 10.1101/2020.01.15.20017236

**Authors:** Shorena Janelidze, Erik Stomrud, Ruben Smith, Sebastian Palmqvist, Niklas Mattsson, David C. Airey, Nicholas K. Proctor, Xiyun Chai, Sergey Shcherbinin, John R. Sims, Jeffrey L. Dage, Oskar Hansson

## Abstract

Cerebrospinal fluid (CSF) p-tau181 (tau phosphorylated at threonine 181) is an established biomarker of Alzheimer’s disease (AD) reflecting abnormal tau metabolism in the brain. Tau can be phosphorylated at multiple other sites including threonine 217, and here we investigated the performance of CSF p-tau217 as a biomarker of AD in comparison to p-tau181. In the Swedish BioFINDER cohort (n=194), p-tau217 had stronger correlations with the tau PET tracer [^18^F]flortaucipir, and more accurately identified individuals with abnormally increased [^18^F]flortaucipir retention. Furthermore, longitudinal increases in p-tau217 were higher compared to p-tau181 and better correlated with [^18^F]flortaucipir retention. P-tau217 correlated better than p-tau181 with PET measures of neocortical amyloid-β burden and more accurately distinguished AD dementia from non-AD neurodegenerative disorders. Higher correlations between p-tau217 and [^18^F]flortaucipir were corroborated in an independent EXPEDITION3 trial cohort (n=32). These findings suggest that p-tau217 might be more useful than p-tau181 in the diagnostic work up of AD.

## INTRODUCTION

Accumulation of intraneuronal neurofibrillary tangles (NFTs) containing paired helical filaments (PHFs) of the microtubule-associated protein tau is one of the defining neuropathological hallmarks of Alzheimer’s disease (AD)^1^. The tau protein has an N-terminal projection domain, a proline-rich region, a repeat region, and a C-terminal domain, with multiple potential phosphorylation sites along all regions^2^. Studies using preparations of PHFs and immunohistochemical staining of postmortem brain tissue with specific tau antibodies established that PHF tau is hyperphosphorylated^3, 4^. High levels of p-tau have consistently been found in cerebrospinal fluid (CSF) of AD patients, and may reflect AD-related tau pathology in the brain^5^. The vast majority of CSF studies have used immunoassays detecting tau phosphorylated at threonine (Thr) 181 (p-tau181)^5^. During the last 2 decades, CSF p-tau181 together with total tau (t-tau) and amyloid-β 42 (Aβ42) have been extensively validated as biomarkers of AD and are currently widely used as diagnostic criteria in research studies, in clinical practice in some countries, and for patient selection in clinical trials^6, 7, 8^. CSF p-tau181 (alone or in combination with Aβ42) accurately differentiates AD from controls and predicts cognitive decline in preclinical and prodromal disease stages^9, 10, 11^. CSF p-tau181 levels are higher in AD compared with other tauopathies including frontotemporal dementia (FTD), progressive supranuclear palsy (PSP) and corticobasal degeneration (CBD) and, hence, CSF p-tau181 has also proven useful in differential diagnosis of dementia^9, 12, 13^.

Postmortem studies have shown that neocortical NFT load and hyperphosphorylated tau immunostaining both correlate with CSF p-tau, although the correlations have been modest^14, 15^. In line with this, recent tau positron emission tomography (PET) investigations have found variable associations between imaging and CSF biomarkers of AD-related tau pathology^16, 17, 18, 19^. The correlations appear to depend on clinical disease stage, since the associations between CSF p-tau181 and tau PET measures were moderate in AD dementia, but weak or absent in cognitively unimpaired individuals and mild cognitive impairment (MCI) patients^18, 19, 20^.

The role of tau phosphorylation at sites other that Thr181 has not been thoroughly investigated with one study demonstrating that p-tau181, p-tau199 and p-tau231 had similar diagnostic accuracies for AD^21^. Recently, CSF levels of another p-tau isoform, p-tau217 (tau phosphorylated at Thr217), was found to correlate with Aβ deposition (measured using [^18^F]-AV45 PET) in AD^22^. Moreover, some preliminary evidence suggest that CSF p-tau217 might correlate more strongly with Tau PET measures than p-tau181 (conference abstract ^23^). Extending these findings from relatively small cohorts, we analyzed CSF p-tau217 using a newly developed ELISA in a total of 226 individuals from two independent cohorts. We tested the associations of p-tau217 with [^18^F]flortaucipir and [^18^F]flutemetamol PET and its diagnostic accuracy for differentiating AD dementia from non-AD neurodegenerative disorders in comparison to p-tau181. Both p-tau isoforms were measured using ELISA kits including the same detection antibody which limited the effects of reagent variability and allowed accurate comparison of biomarker performance.

## RESULTS

### Participants

We included 65 cognitively unimpaired controls (CU), 29 Aβ^+^ MCI (MCI due to AD [MCI-AD]), 43 AD dementia patients, and 57 patients with non-AD neurodegenerative disorders (Table 1). There were no differences in sex and education between different diagnostic groups, but patients with non-AD neurodegenerative disorders were somewhat younger on average than CU. The core CSF AD biomarkers (Aβ42, Aβ42/Aβ40, p-tau181) were increasingly abnormal in MCI-AD and AD dementia.

**Table 1.** Demographic and clinical characteristics.

|  | CU<br>n=65 | MCI-AD<br>n=29 | AD<br>n=43 | Non-AD disorders <sup>*</sup><br>n=57 |
| --- | --- | --- | --- | --- |
| Age, years | 75 (7) | 72 (10) | 72 (7) | 70 (6) <sup>a</sup> |
| Sex F/M, n | 33/32 | 10/19 | 20/23 | 23/34 |
| Education, years** | 12 (4) | 12 (4) | 12 (4) | 13 (4) |
| MMSE** | 29 (1) | 26 (3) <sup>a</sup> | 21 (5) <sup>a</sup> | 24 (6) <sup>a</sup> |
| Aβ42, pg/ml | 508 (186) | 337 (110) <sup>a</sup> | 299 (100) <sup>a</sup> | 526 (251) |
| Aβ42/Aβ40 | 0.09 (0.04) | 0.05 (0.01) <sup>a</sup> | 0.06 (0.02) <sup>a</sup> | 0.12 (0.04) <sup>a</sup> |
| p-tau217, pg/ml | 188 (148) | 684 (388) <sup>a</sup> | 812 (570) <sup>a</sup> | 135 (199) <sup>b</sup> |
| p-tau181, pg/ml | 153 (86) | 373 (167) <sup>a</sup> | 392 (221) <sup>a</sup> | 111 (89) <sup>a</sup> |
| Aβ positivity*** | 61% | 100% | 100% | 53% |

Data are shown as mean (SD) unless otherwise specified. Differences between the groups were tested using Mann-Whitney *U* and chi-square tests. <sup>a</sup> $p < 0.001$ , <sup>b</sup> $p < 0.01$ .
\* Non-AD neurodegenerative disorders group included 10 PD, 17 PDD, 6 PSP, 7 DLB, 7 CBS, 4 SD and 6 bvFTD patients.
\*\* Education is missing for 1 CU and 2 FTD patients; MMSE is missing for 1 CU and 1 MCI-AD, 3 AD dementia and 1 FTD patients.
\*\*\* Aβ positivity by either CSF Aβ 42 or [<sup>18</sup>F]flutemetamol PET.
AD, Alzheimer's disease dementia; bvFTD, behavioral-variant frontotemporal dementia; CBS, corticobasal syndrome; CU, cognitively unimpaired; DLB, dementia with Lewy bodies; F, female; M, male; MCI-AD, mild cognitive impairment due to Alzheimer's disease; MMSE, Mini Mental State Examinations; PD, Parkinson's disease; PDD, Parkinson's disease with dementia; PSP, progressive supranuclear palsy; SD semantic dementia.

### Group comparisons

CSF p-tau217 and p-tau181 were highly correlated (whole cohort Pearson r=0.973; CU Pearson r=0.979; MCI-AD Pearson r=0.972; AD dementia Pearson r=0.958; non-AD neurodegenerative disorders Pearson r=0.969; all p<0.001). Both biomarkers as well as their ratios to t-tau were increased in the 3 Aβ^+^ groups compared to Aβ^-^ CU and non-AD groups (Fig. 1). At the same time, the dynamic range was greater for p-tau217: the mean CSF concentration of p-tau217 was 7.3-8.6 fold higher in Aβ^+^ MCI and Aβ^+^ AD than in Aβ^-^ CU, whereas we only observed a 3.6-3.7 fold increase for p-tau181.

**Figure 1.**
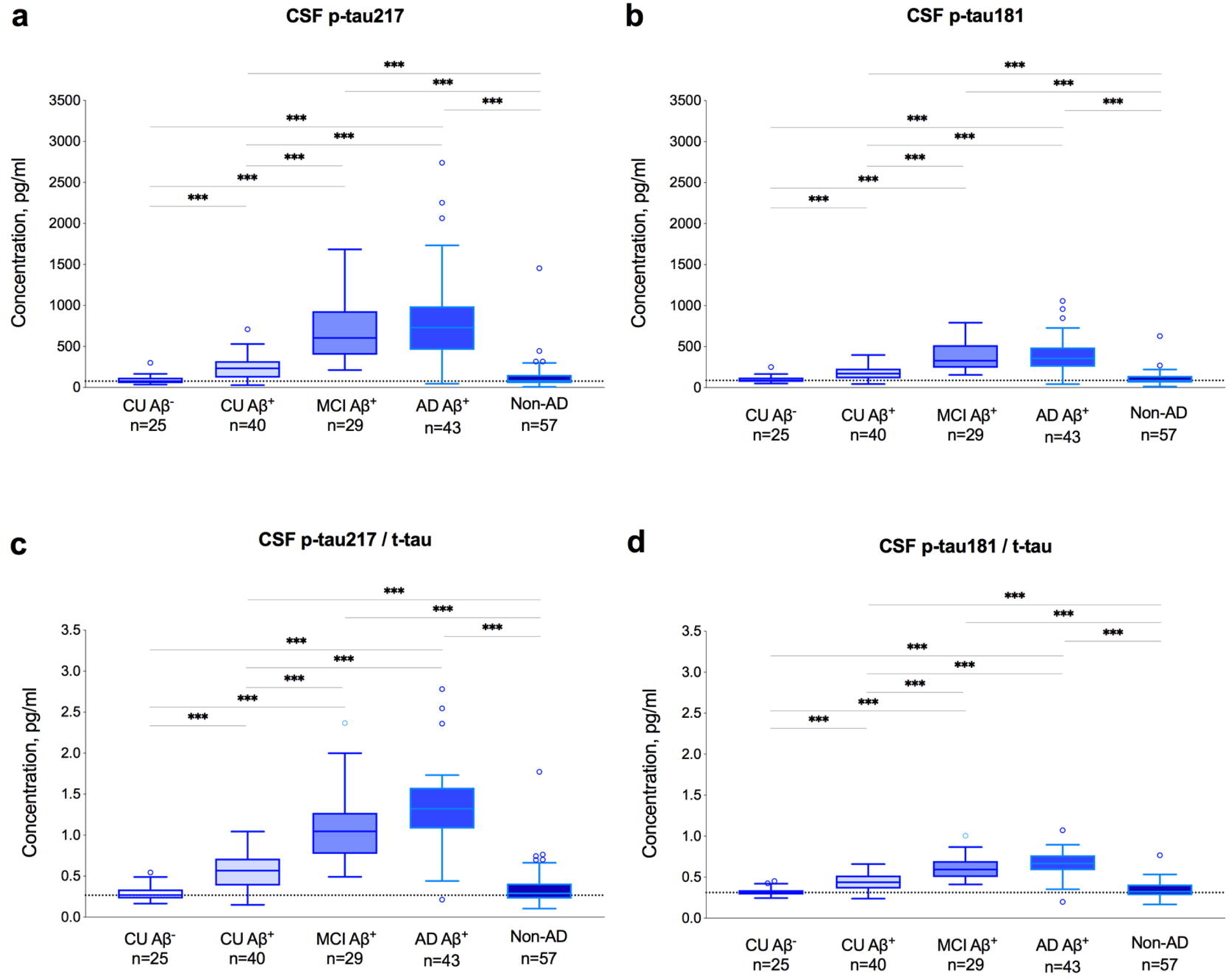
CSF p-tau in diagnostic groups. CSF p-tau217 (**a**), p-tau181 (**b**), CSF p-tau217/t-tau (**c**) and CSF p-tau181/t-tau (**d**) in CU Aβ^+^, CU Aβ^+^, MCI Aβ^+^, AD Aβ^+^ and non-AD neurodegenerative disorders. Non-AD neurodegenerative disorders group included 10 PD, 17 PDD, 6 PSP, 7 DLB, 7 CBS, 4 SD and 6 bvFTD patients. The dotted lines indicate median levels in the CU Aβ^-^ group. P values are from general linear models adjusted for age and sex. Abbreviations: AD, Alzheimer’s disease; bvFTD, behavioral-variant frontotemporal dementia; CBS, corticobasal syndrome; CSF, cerebrospinal fluid; CU, cognitively unimpaired controls; DLB, dementia with Lewy bodies; MCI, Mild Cognitive Impairment; PD, Parkinson’s disease; PDD, Parkinson’s disease with dementia; PSP, progressive supranuclear palsy; SD semantic dementia.

### Associations with [^18^F]flortaucipir and [^18^F]flutemetamol

[^18^F]flortaucipir PET was performed in 184 study participants. First, we studied associations between the two p-tau isoforms and [^18^F]flortaucipir retention in four commonly used pre-defined brain regions including inferior temporal cortex and composites corresponding to Braak I/II, III/IV and V/VI regions of interest (ROIs)^24^. We found that higher levels of p-tau217 and p-tau181 were both associated with increased [^18^F]flortaucipir retention in all four regions (Fig. 2 and Table 2). However, correlation coefficients were consistently higher for p-tau217 compared with p-tau181 and the differences between the coefficients were statistically significant (Fig. 2 and Table 2). Next, we studied the associations between CSF p-tau and [^18^F]flortaucipir in CU, MCI-AD and AD dementia cases, separately. In CU, both p-tau isoforms were significantly related to [^18^F]flortaucipir retention in the earliest tau region (the Braak I/II ROI) but not in the three other regions, whereas in AD dementia the correlations were significant in all regions except the Braak I/II ROI (Table 2). Furthermore, in MCI-AD, p-tau181 correlated significantly with [^18^F]flortaucipir uptake only in the Braak I/II ROI while p-tau217 correlated in all four regions. Importantly, the correlation coefficients were higher for [^18^F]flortaucipir and CSF p-tau217 compared to CSF p-tau181 in all three diagnostic groups and across all regions (Fig. 2 and Table 2).

**Figure 2.**
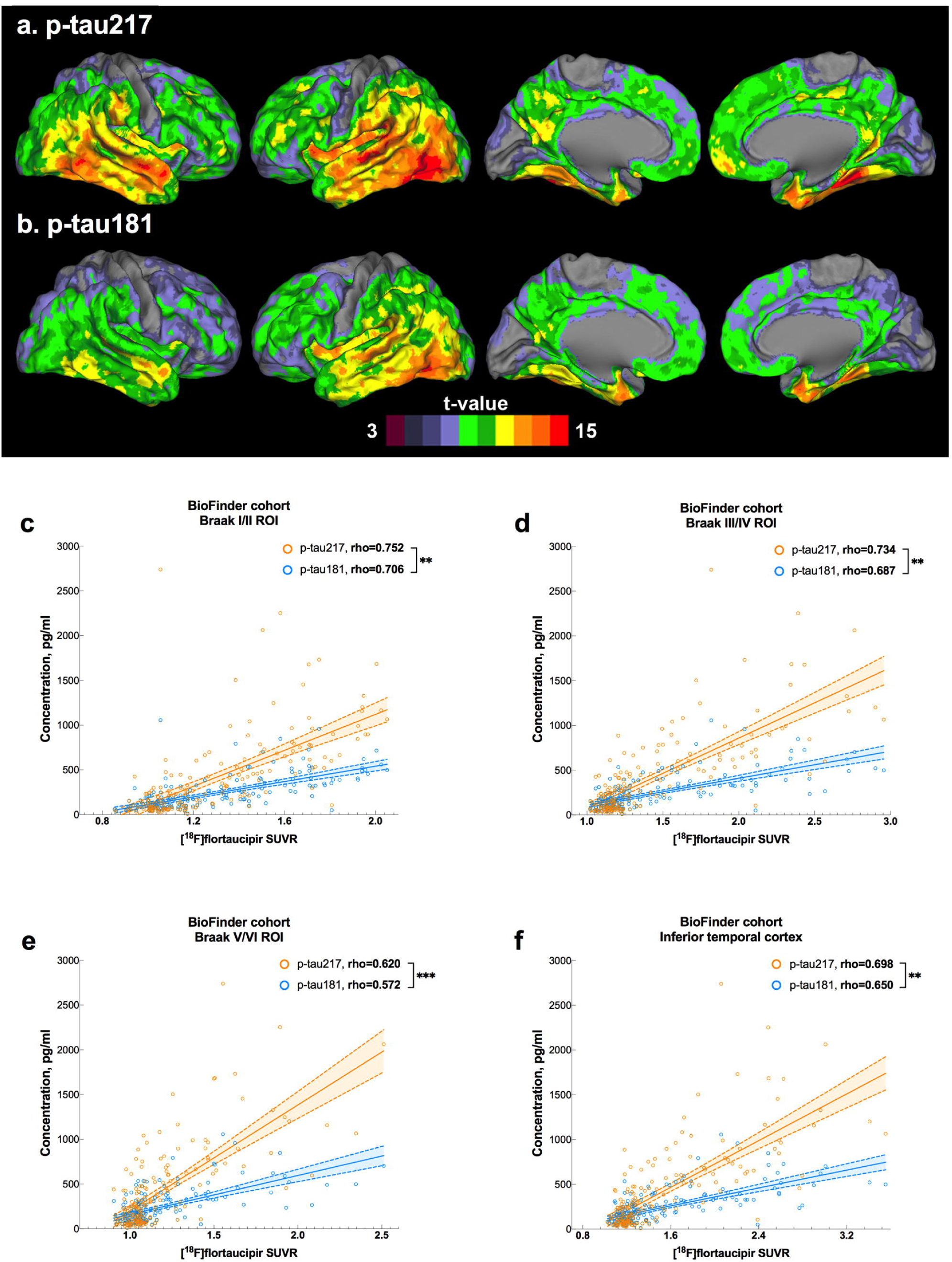
Associations between[^18^F]flortaucipir and p-tau in the BioFINDER cohort. Voxel-wise regression analysis of p-tau217 (**a**) and p-tau181 (**b**) vs [^18^F]flortaucipir corrected for age. Associations between[^18^F]flortaucipir retention in *a priori* defined brain regions linked to tau pathology in AD and CSF p-tau217 and p-tau181 (**c-f**). Data are shown as Spearman correlation coefficients (rho). Differences between the correlation coefficients were tested using estimated Spearman coefficients and method described in Rosner et al.^63^. Abbreviations: ROI, region of interest; SUVR, standardized uptake value ratio.

**Table 2.**
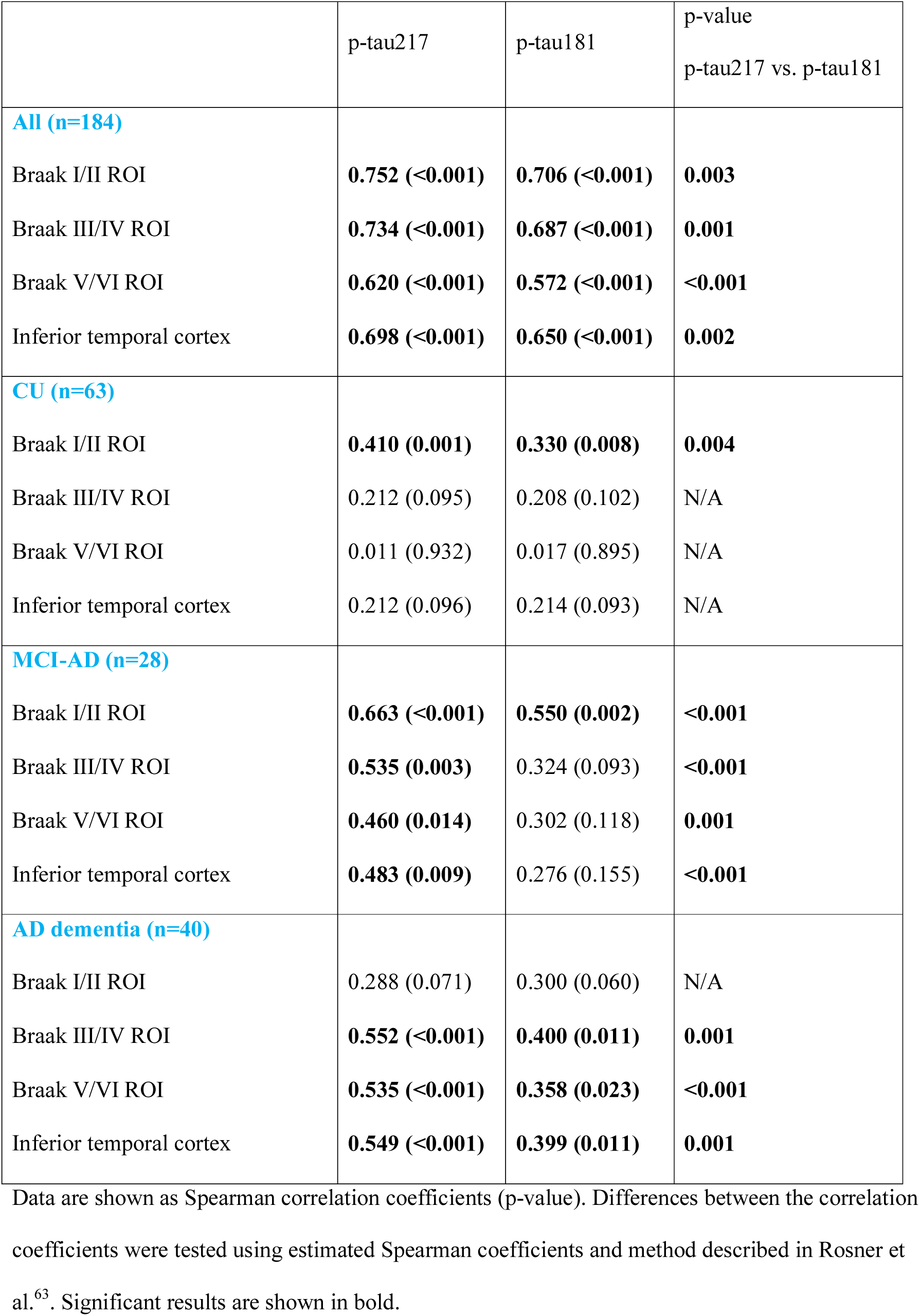
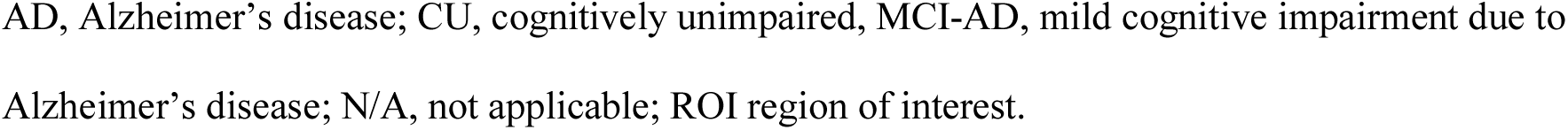
Spearman correlations between CSF p-tau and regional [^18^F]flortaucipir.

|  | p-tau217 | p-tau181 | p-value<br>p-tau217 vs. p-tau181 |
| --- | --- | --- | --- |
| **All (n=184)** |  |  |  |
| Braak I/II ROI | **0.752 (<0.001)** | **0.706 (<0.001)** | **0.003** |
| Braak III/IV ROI | **0.734 (<0.001)** | **0.687 (<0.001)** | **0.001** |
| Braak V/VI ROI | **0.620 (<0.001)** | **0.572 (<0.001)** | **<0.001** |
| Inferior temporal cortex | **0.698 (<0.001)** | **0.650 (<0.001)** | **0.002** |
| **CU (n=63)** |  |  |  |
| Braak I/II ROI | **0.410 (0.001)** | **0.330 (0.008)** | **0.004** |
| Braak III/IV ROI | 0.212 (0.095) | 0.208 (0.102) | N/A |
| Braak V/VI ROI | 0.011 (0.932) | 0.017 (0.895) | N/A |
| Inferior temporal cortex | 0.212 (0.096) | 0.214 (0.093) | N/A |
| **MCI-AD (n=28)** |  |  |  |
| Braak I/II ROI | **0.663 (<0.001)** | **0.550 (0.002)** | **<0.001** |
| Braak III/IV ROI | **0.535 (0.003)** | 0.324 (0.093) | **<0.001** |
| Braak V/VI ROI | **0.460 (0.014)** | 0.302 (0.118) | **0.001** |
| Inferior temporal cortex | **0.483 (0.009)** | 0.276 (0.155) | **<0.001** |
| **AD dementia (n=40)** |  |  |  |
| Braak I/II ROI | 0.288 (0.071) | 0.300 (0.060) | N/A |
| Braak III/IV ROI | **0.552 (<0.001)** | **0.400 (0.011)** | **0.001** |
| Braak V/VI ROI | **0.535 (<0.001)** | **0.358 (0.023)** | **<0.001** |
| Inferior temporal cortex | **0.549 (<0.001)** | **0.399 (0.011)** | **0.001** |

Data are shown as Spearman correlation coefficients (p-value). Differences between the correlation coefficients were tested using estimated Spearman coefficients and method described in Rosner et al.<sup>63</sup>. Significant results are shown in bold.
AD, Alzheimer's disease; CU, cognitively unimpaired, MCI-AD, mild cognitive impairment due to Alzheimer's disease; N/A, not applicable; ROI region of interest.

We also examined the correlations between the CSF p-tau to t-tau ratios with [^18^F]flortaucipir. In the AD dementia group, we observed that correlation coefficients were considerably higher for the p-tau217/t-tau ratio (rho=0.742-0.768) than for p-tau217 alone (rho=0.535-0.552) whereas the correlation were comparable for p-tau181/t-tau ratio (rho=0.358-0.400) and for p-tau181 (rho=0.318-0.420) (Supplementary Table 1).

To be able to establish whether CSF p-tau could predict normal vs abnormal Tau PET scans we dichotomized [^18^F]flortaucipir outcomes based on the previously established SUVR cutoffs of 1.3^25^. Compared to p-tau181, p-tau217 was a significantly more accurate predictor of abnormal Tau PET status in all 4 regions (Table 3).

**Table 3.** Receiver Operating Characteristic (ROC) analysis of CSF p-tau217 and p-tau181 for identifying abnormal ^18^F[Flortaucipir status and differentiating AD dementia from a group of non-AD neurodegenerative disorders.

|  | AUC | Cut-off<br>pg/ml | Sensitivity<br>% | Specificity<br>% | Youden's J |
| --- | --- | --- | --- | --- | --- |
| **Braak I/II ROI** |  |  |  |  |  |
| p-tau217 | 0.933<sup>a</sup> (0.898-0.968) | 287.8 | 88 | 87 | 0.741 |
| p-tau181 | 0.912 (0.869-0.955) | 217.0 | 82 | 88 | 0.694 |
| **Braak III/IV ROI** |  |  |  |  |  |
| p-tau217 | 0.933<sup>b</sup> (0.896-0.970) | 441.6 | 80 | 94 | 0.744 |
| p-tau181 | 0.900 (0.851-0.949) | 224.7 | 82 | 86 | 0.674 |
| **Braak V/VI ROI** |  |  |  |  |  |
| p-tau217 | 0.923<sup>b</sup> (0.874-0.973) | 582.9 | 87 | 88 | 0.747 |
| p-tau181 | 0.876 (0.805-0.947) | 231.6 | 94 | 73 | 0.668 |
| **Inferior temporal** |  |  |  |  |  |
| p-tau217 | 0.886<sup>b</sup> (0.833-0.938) | 418.0 | 74 | 93 | 0.663 |
| p-tau181 | 0.855 (0.795-0.915) | 219.4 | 75 | 86 | 0.611 |
| **AD dementia vs** |  |  |  |  |  |
| p-tau217 | 0.943<sup>c</sup> (0.892-0.994) | 246.1 | 91 | 91 | 0.819 |
| p-tau181 | 0.914 (0.848-0.980) | 235.3 | 79 | 96 | 0.756 |

Data are shown as AUC (95% CI). [ $^{18}\text{F}$ ]flortaucipir data was dichotomized based on the SUVR cutoff of 1.3<sup>25</sup>. <sup>a</sup>p=0.010 compared with p-tau181, <sup>b</sup>p<0.001 compared with p-tau181, <sup>c</sup>p=0.026 compared with p-tau181.
AUC, area under the curve; CI, confidence interval; ROI, region of interest.

In a subcohort of study participants who underwent [^18^F]flutemetamol PET (n=139), we found that high levels of p-tau217 and p-tau181 were associated with increased neocortical [^18^F]flutemetamol retention and that the correlations were stronger for p-tau217 compared with p-tau181 (rho=0.655 vs. rho=0.601, p=0.002). We corroborated these findings in a previously described cohort of CU and MCI patients (n=330)^26^ from the Swedish BioFINDER study where correlations with composite [^18^F]flutemetamol SUVR were again stronger for p-tau217 than p-tau181 (rho=0.666 vs. rho=0.612, p=0.003).

### Diagnostic accuracy of CSF p-tau217 and p-tau181 for AD dementia vs non-AD neurodegenerative disorders

Next, we investigated diagnostic accuracies of CSF p-tau217 and p-tau181 in differentiating AD dementia (n=43) from non-AD neurodegenerative disorders (n=57). In ROC analysis, p-tau217 showed significantly larger AUC than p-tau181 (0.943 vs. 0.914, p=0.026; Table 3). When using Youden index derived cutoffs, the sensitivity of p-tau217 was 12% higher (91% vs. 79%), whereas specificity was 5% lower (91% vs 96%) compared with p-tau181. However, for cutoffs with a fixed sensitivity of 91%, p-tau217 showed higher specificity than p-tau181 (91% vs. 75%).

### Associations between longitudinal changes in CSF p-tau and [^18^F]flortaucipir

Ninety-seven study participants underwent additional earlier LP before the main LP (which was more close to [^18^F]flortaucipir PET). In these individuals, we analyzed p-tau in two CSF samples and assessed associations between [^18^F]flortaucipir retention and the annual rate of change in the biomarker levels. The mean time between the two LPs was 3.4 years (SD=1.4). We found larger annualized increase in p-tau217 (21.0pg/ml [44.9], mean [SD]) than p-tau181 (9.4pg/ml [16.2], mean [SD]). Furthermore, longitudinal increases in the levels of p-tau217 and p-tau181 were associated with higher regional [^18^F]flortaucipir uptake. These correlations were significant only in the Aβ^+^ group and higher for p-tau217 than for p-tau181 (Table 4). In addition, for exploratory group-wise analyses, we compared longitudinal changes in the CSF p-tau levels between individuals that were [^18^F]flortaucipir negative or had an abnormal uptake only in the Braak I/II ROI (Braak 0/I/II^+^ group) vs. study participants with abnormal [^18^F]flortaucipir retention in the Braak III/IV and V/VI ROIs (Braak III/IV^+^ and V/VI^+^ group) using the SUVR cutoff of 1.3^25^ (Fig. 3). The rates of yearly increase in the levels of p-tau 217 were higher in both Braak III/IV^+^ and V/VI^+^ groups compared to 0/I/II^+^ group, but also in the Braak III/IV^+^ compared to V/VI^+^ groups. However, for p-tau181, the increase was only higher in the latest ROI V/VI^+^ group compared to the 0/I/II^+^ group with no other significant differences between the groups.

**Table 4.** Spearman correlations between regional [^18^F]flortaucipir and longitudinal changes CSF p-tau.

| | $\Delta\text{p-tau217}$ | $\Delta\text{p-tau181}$ | p-value<br>$\Delta\text{p-tau217}$ vs. $\Delta\text{p-tau181}$ |
| --- | --- | --- | --- |
| **All (n=97)** |  |  |  |
| Braak I/II ROI | **0.507 (<0.001)** | **0.381 (<0.001)** | **0.001** |
| Braak III/IV ROI | **0.456 (<0.001)** | **0.339 (0.001)** | **0.012** |
| Braak V/VI ROI | **0.311 (0.002)** | **0.201 (0.048)** | **0.011** |
| Inferior temporal cortex | **0.455 (<0.001)** | **0.358 (<0.001)** | **0.036** |
| **Aβ positive (n=67)** |  |  |  |
| Braak I/II ROI | **0.519 (0.001)** | **0.376 (0.002)** | **0.004** |
| Braak III/IV ROI | **0.505 (<0.001)** | **0.335 (0.006)** | **<0.001** |
| Braak V/VI ROI | **0.369 (0.002)** | 0.189 (0.126) | **<0.001** |
| Inferior temporal cortex | **0.489 (<0.001)** | **0.329 (0.007)** | **<0.001** |
| **Aβ negative (n=30)** |  |  |  |
| Braak I/II ROI | 0.329 (0.075) | 0.151 (0.426) | N/A |
| Braak III/IV ROI | -0.044 (0.816) | 0.160 (0.398) | N/A |
| Braak V/VI ROI | -0.119 (0.533) | 0.081 (0.670) | N/A |
| Inferior temporal cortex | 0.003 (0.988) | 0.276 (0.140) | N/A |

Delta ( $\Delta$ ) p-tau217 and $\Delta\text{p-tau181}$ represent yearly changes in the biomarker levels. Data are shown as Spearman correlation coefficients (p-value). Differences between the correlation coefficients were tested using estimated Spearman coefficients and method described in Rosner et al.<sup>63</sup>.
Significant results are shown in bold.
Aβ , amyloid-β ; N/A, not applicable; ROI, region of interest.

**Figure 3.**
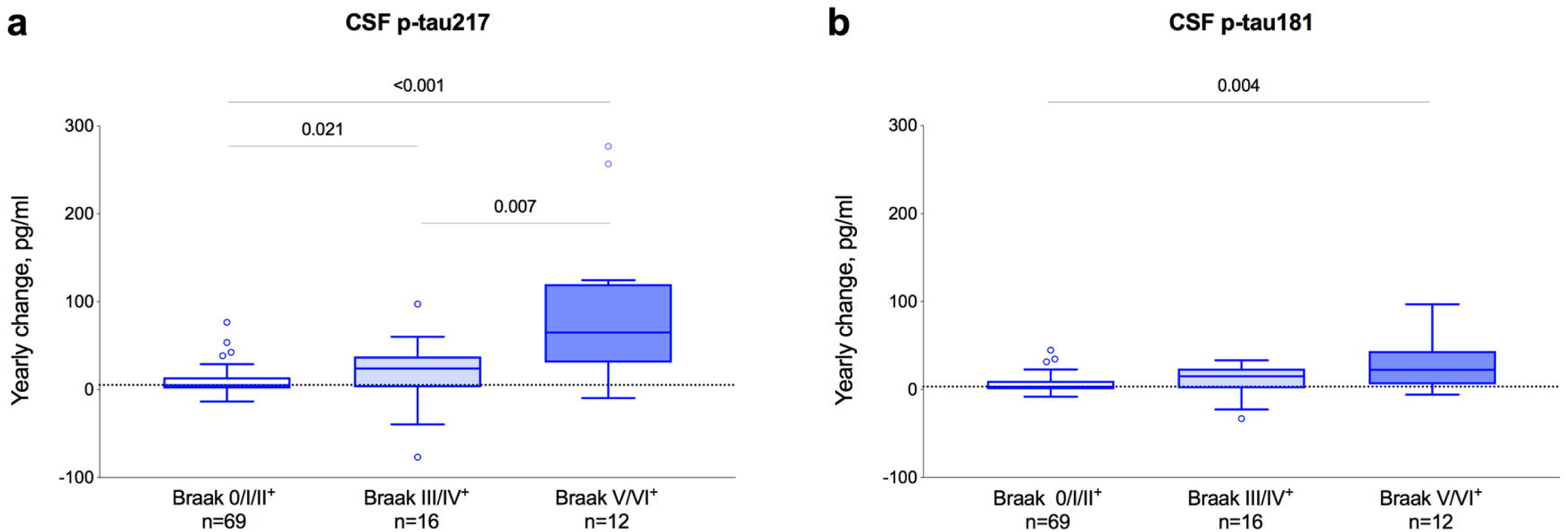
Longitudinal changes in CSF p-tau across the Braak ROI groups. Study participants were staged into different Braak ROI groups using [^18^F]flortaucipir PET. [^18^F]flortaucipir data was dichotomized based on the SUVR cutoff of 1.3^25^. Annual changes in CSF p-tau217 (**a**) and p-tau181 (**b**) in the Braak 0/I/II (normal [^18^F]flortaucipir retention or abnormal [^18^F]flortaucipir retention limited to ROI I/II), III/IV (abnormal [^18^F]flortaucipir retention in ROIs III/IV) and V/VI (abnormal [^18^F]flortaucipir retention in ROI V/VI) groups. The dotted lines indicate median levels in the Braak 0/I/II group. P values are from Mann-Whitney test. Abbreviations: CSF, cerebrospinal fluid; ROI, region of interest.

### Associations of CSF p-tau217 and p-tau181 with [^18^F]flortaucipir in an independent validation cohort

To validate observations in the BioFINDER cohort, relationships between the two p-tau isoforms and [^18^F]flortaucipir signal were examined for amyloid positive mild AD participants of the EXPEDITION3 trial (n=32, Supplementary Table 2). Although the population, PET acquisition and processing parameters differ, we found close similarity between findings in the BioFINDER and EXPEDITION3 cohorts. Specifically, we found a positive association between levels of p-tau217 and p-tau181 and flortaucipir SUVR (Fig. 4). Moreover, the correlation coefficient was higher for p-tau217 compared with p-tau181.

**Figure 4.**
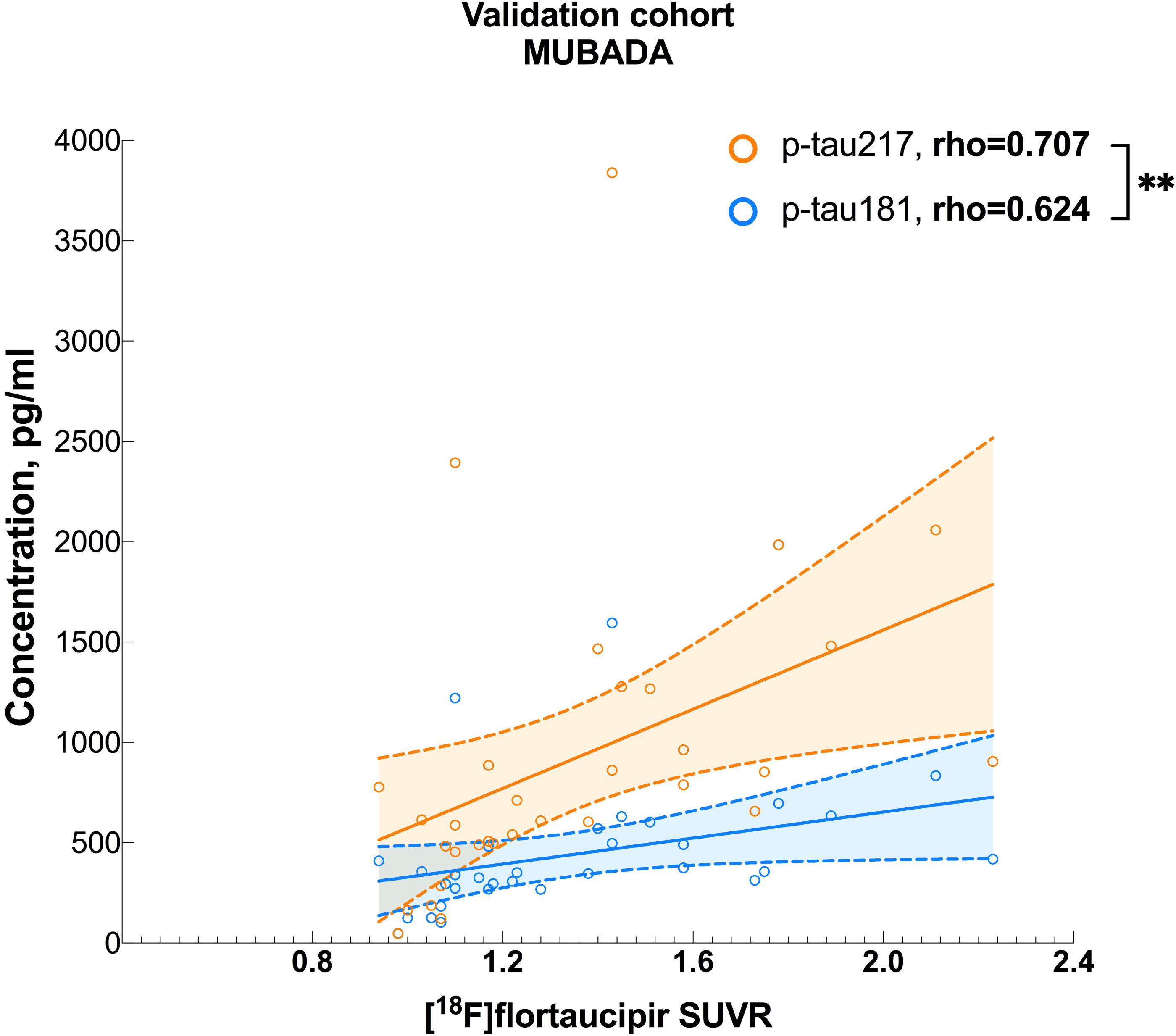
Associations between[^18^F]flortaucipir and p-tau in the validation cohort. Associations between[^18^F]flortaucipir MUBADA SUVR and CSF p-tau217 and p-tau181. Data are shown as Spearman correlation coefficients (rho). Differences between the correlation coefficients were tested using estimated Spearman coefficients and method described in Rosner et al.^63^. Abbreviations: SUVR, standardized uptake value ratio.

## DISCUSSION

Here we present evidence that CSF p-tau217 is a better biomarker of AD pathology than CSF p-tau181. CSF concentrations of p-tau217 in prodromal AD and AD dementia were several fold higher compared with p-tau181. CSF p-tau217 showed higher correlations with [^18^F]flortaucipir and [^18^F]flutemetamol retention. Furthermore, this biomarker more accurately identified individuals with abnormally increased [^18^F]flortaucipir binding in different neocortical regions linked to tau pathology in AD. In addition, p-tau217 was significantly better than p-tau181 in distinguishing AD from a group of other neurodegenerative disorders. Longitudinal increases in p-tau217 were higher compared to p-tau181 and better correlated with [^18^F]flortaucipir retention in Aβ^+^ individuals. Finally, the findings of higher correlations between [^18^F]flortaucipir and p-tau217 compared with p-tau181 were validated in an independent cohort.

[^18^F]flortaucipir was developed to detect PHF-tau^27^ with several autoradiography investigations confirming its selective binding to tau aggregates containing PHF in AD^28, 29, 30^. *In vivo* retention of [^18^F]flortaucipir has been demonstrated to strongly correlate with the density of intrasomal tau tangles and tau-positive neurites in AD postmortem brain tissue^31^. Thr217 is one of roughly 40 different phosphorylation sites identified in PHF tau from Alzheimer brain^3^. Several antibodies specific for p-tau have been commonly used for immunostaining of human brain section to visualize and quantify tau pathology. One of these antibodies, AT100, binds to a conformational epitope that requires dual phosphorylation at Thr212 and serine (Ser) 214^32^. However, some data suggests that phosphorylation of nearby Thr217 is necessary for secondary phosphorylation of Thr212 and Ser 214 and that Thr217 is a part of A100 epitope^33, 34^. Interestingly, AT270, an antibody that recognizes p-tau181, has been shown to stain fewer NFTs and neuropil threads in brain tissue from AD patients than AT100 ^35^, indicating that CSF p-tau217 might better reflect the pathological state of tau associated with PHF-tau formation. Accordingly, we found that correlations with [^18^F]flortaucipir in different brain region were consistently higher for p-tau217 than p-tau181 and that [^18^F]flortaucipir retention was more related to longitudinal changes in p-tau217 than in p-tau181. In line with previous studies, the relationships between the CSF biomarkers and [^18^F]flortaucipir were disease stage dependent^16, 17, 18, 19, 20^. In the CU and MCI-AD groups, higher levels of both CSF p-tau isoforms were associated with increased [^18^F]flortaucipir retention in the Braak I/II ROI (entorhinal cortex), one of earliest region of tau pathology according to the Braak staging system^36, 37^. In patients with AD dementia, these associations were only significant in more late Braak III/IV and V/VI ROIs and inferior temporal cortex. Importantly, we also observed significant correlations between p-tau217 and [^18^F]flortaucipir uptake in the Braak III/IV and V/VI ROIs and inferior temporal cortex in the MCI-AD group where the correlations were not significant for p-tau181. Furthermore, p-tau217 was superior in detecting pathological [^18^F]flortaucipir status in all four regions. Thus, our results suggest that CSF p-tau217 better mirrors PET measures of tau pathology in different AD stages and especially in prodromal AD. Increase in CSF p-tau181 is considered to be specific for AD^38^. CSF levels of this biomarker have been shown to associate with Aβ PET^39^ and improve differentiation of AD from other tauopathies^9, 12, 13^. Providing further support for advantages of using p-tau217 over p-tau181 as a biomarker of AD, we found that CSF levels of p-tau217 correlated better with [^18^F]flutemetamol retention and distinguished more accurately AD from other dementias.

CSF p-tau181 is one of the core biomarkers incorporated into the NIA-AA Research Framework to define AD^38^. There is an ongoing effort to standardize protocols for determination of biomarker levels and to develop criteria for appropriate use of CSF testing in the diagnosis of AD with an intent of implementing CSF biomarkers into clinical practice^40, 41^. The findings of the present study suggest that CSF p-tau217 is consistently more strongly related to the AD pathological process and might be more useful than p-tau181 in the diagnostic work up of AD. However, future validation studies in independent cohorts are required before adopting CSF p-tau217 as a clinically relevant biomarker.

## METHODS

The BioFINDER study was approved by the Regional Ethics Committee in Lund, Sweden, and all participants gave their written informed consent to participate in the study. Samples for the validation cohort were collected as part of a phase 3 clinical trial conducted in accordance with the Declaration of Helsinki for experiments involving human research. All participants gave their informed consent to participate in the study.

### Study population

#### BioFINDER cohort

Cognitively unimpaired controls (n=65) and patients with MCI^42^ (n=29), AD dementia^43^ (n=43), PD^44^ (n=10), PDD^45^ (n=17), PSP^46^ (n=6), DLB^47^ (n=7), CBS^48^ (n=7) and FTD^49, 50^ (n=10) were recruited from the Swedish BioFINDER study. All participants underwent a medical history, complete neurologic examination, neuropsychological testing and LP. Two LPs were performed in 97 study participants. One hundred eighty-four individuals (63 CU, 28 MCI, 70 AD, 10 PD, 16 PDD, 6 PSP, 5 DLB, 7 CBS and 9 FTD) underwent [^18^F]flortaucipir PET. [^18^F]flutemetamol PET was performed in 139 study participants (61 CU, 24 MCI, 37 AD, 3 PSP, 3 DLB, 5 CBS and 6 FTD). In this study, all MCI and AD patients as well as 39 (61%) CU were Aβ positive (according to either CSF Aβ42 or [^18^F]flutemetamol PET). CU included 55 cognitively healthy individuals and 10 participants with subjective cognitive decline. Recruitment and inclusion criteria for CU have been previously described^51^. The characteristics of the study participants are given in Table 1.

#### Validation cohort

We validated our observations using baseline biomarker storage samples and data acquired in EXPEDITION3 phase 3 trial with solanezumab (NCT01900665). EXPEDITION3 was a double-blind, placebo-controlled, phase 3 trial involving Aβ positive (shown by means of florbetapir PET or Aβ1-42 measurements in CSF) patients with mild dementia due to AD, defined as a Mini–Mental State Examination (MMSE) score of 20 to 26^52^. Tau scanning with [^18^F]flortaucipir was performed at baseline, 40 weeks, and 80 weeks in subset of subjects (approximately 10% of total subjects). CSF samples were collected during the trial in a subset of subjects and aliquots stored for future biomarker research. Overall, 32 participants had undergone both flortaucipir PET and lumbar puncture at baseline (before any treatment) and comprised the validation cohort. The characteristics of those participants are given in Supplementary Table 2.

### CSF sampling and analysis

The procedure and analysis of CSF followed the Alzheimer’s Association Flow Chart for CSF biomarkers^9^. Lumbar CSF samples were collected at the 2 centers and analyzed according to a standardized protocol^53^. CSF Aβ42 and t-tau were measured with EUROIMMUN kits and according to the manufacturer’s recommendations.

#### CSF p-tau181 and p-tau217 measurements

The p-tau181 and p-tau217 assays were designed for CSF analysis. Both assays were performed on a streptavidin small spot plate using the Meso Scale Discovery (MSD) platform (Meso Scale Discovery, Rockville, MD, USA). Anti-p-tau217 antibody IBA413 was used as a capture antibody in the p-tau217 assay whereas anti-p-tau181 antibody AT270 was used as a capture antibody in the p-tau181 assay. Antibodies were conjugated with Biotin (Thermo Scientific, catalog number: 21329) or SULFO-TAG (MSD, catalog number: R91AO-1). The assays were calibrated using a recombinant tau (4R2N) protein that was phosphorylated in vitro using a reaction with glycogen synthase kinase-3 and characterized by mass spectrometry. The samples were thawed on wet ice, briefly vortexed, and diluted 1:8 in Diluent 35 (MSD, catalog number: R50AE) with the addition of a heterophilic blocking reagent to a concentration of 200µg/ml (Scantibodies Inc, catalog number: 3KC533). In order to perform the assays, MSD small-spot streptavidin coated plates (MSD, catalog number: L45SA) were blocked for 1 hour at room temperature with 200µl of 3% BSA in DPBS with 650rpm shaking on a plate shaker. The plates were then washed three times with 200µl of wash buffer (PBS + 0.05% Tween 20) and 25µl of biotinylated capture antibody (AT270 for p-tau181 or IBA413 for p-tau217) at 1µg/ml and 0.1 µg/ml respectively were added to the wells and incubated for 1 hour at room temperature with 650rpm shaking on a plate shaker. The plates were again washed three times with 200µl of wash buffer and 50µl of diluted calibrator or sample was added to each well and incubated for 2 hours at room temperature with 650rpm shaking on a plate shaker. The plates were then washed three times with 200µl of wash buffer and 25µl of SULFO-tagged LRL detection antibody was added at 3µg/ml for the p-tau181 and at 0.5µg/ml for the p-tau217 plates and incubated for 1 hour at room temperature with 650rpm shaking on a plate shaker. The plates were washed a final time with 200µl of wash buffer and 150µl of 2x MSD Read Buffer T with Surfactant (MSD, catalog number: R92TC) was added to each plate and read on the MSD SQ120 within 10 minutes of read buffer addition. Samples were analyzed in duplicates and the mean of duplicates were used in statistical analysis. The assay performances across all p-tau217 and p-tau181 plates are summarized in Supplementary Table 3 with a 10pg/ml and 50pg/ml QC buffer spike and high, medium, and low control CSF samples. Individual measurements all fell within 20% of the mean for QC and control samples.

### Tau and Aβ PET imaging and processing

#### BioFINDER cohort

[^18^F]flortaucipir and [^18^F]flutemetamol were synthesized at Skåne University Hospital, Lund, and PET scans were performed on a GE Discovery 690 PET scanner (Flortaucipir; General Electric Medical Systems, Chicago, IL) and a Philips Gemini TF 16 scanner (Flutemetamol; Philips Healthcare, Amsterdam, the Netherlands), respectively, as previously described^24, 54^.

The mean injected dose of [^18^F]flortaucipir was ≈370MBq, and participants underwent a PET scan during the 80-to 100-minute interval after injection. Images were motion corrected with the AFNI 3dvolreg, time averaged, and rigidly coregistered to the skull-stripped MRI scan. Standardized uptake value ratio (SUVR) images were created using inferior cerebellar gray matter as reference region^55^. FreeSurfer (version 5.3) parcellation of the T1-weighted MRI scan was applied to the PET data transformed to participants’ native T1 space to extract mean regional SUVR values for each participant in four predefined ROI including inferior temporal cortex and three regions corresponding to different image[based stages of tau as described in Cho et al.^56^: the Braak I/II (entorhinal cortex), III/IV (parahippocampal gyrus, fusiform gyrus, amygdala, inferior temporal and middle temporal gyri) and V/VI (posterior cingulate gyrus, caudal anterior cingulate gyrus, rostral anterior cingulate gyrus, precuneus, inferior parietal lobule, superior parietal lobule, insula, supramarginal gyrus, lingual gyrus, superior temporal gyrus, medial orbitofrontal gyrus, rostral middle frontal gyrus, lateral orbitofrontal gyrus, caudal middle frontal gyrus, superior frontal gyrus, lateral occipital gyrus, precentral gyrus, postcentral gyrus and paracentral gyrus) ROIs. For voxel-wise analysis between [^18^F]flortaucipir and CSF p-tau the MR and PET images were transformed into Montreal Neurological Institute space (2mm MNI152 MRI template) and voxel-wise correlations were made using multiple regressions adjusting for age in SPM12 (http://www.fil.ion.ucl.ac.uk/spm). Images were thresholded using family-wise error (FWE) correction at p<0.01. The thresholded images were overlaid on a Population-Average, Landmark- and Surface-based (PALS) image ^57^ using CARET v5.65 (Van Essen Lab; http://brainvis.wustl.edu). The images were not corrected for partial volume effects.

For [^18^F]flutemetamol, the mean injected dose was ≈185 MBq. PET images were acquired between 90 and 110 minutes after injection. The scanning and processing procedures have been described previously^53, 58^. The weighted mean standardized uptake value ratio (SUVR) from a global neocortical region of interest was calculated relative to a composite reference region (white matter, cerebellum and brainstem)^58, 59^.

#### Validation cohort

The tau PET acquisitions were performed from 75 to 105 minutes (6 x 5 min frames) after injection of approximately 240 MBq of [^18^F]flortaucipir. Frames were aligned and averaged with an acquisition time-offset correction. Average 75-105 min image was spatially registered to the corresponding individual subject’s MRI space and then to the MRI template in Montreal Neurological Institute (MNI) stereotaxic space. Reference signal was parametrically derived in the white matter-based region to isolate non-specific signal (parametric estimate of reference signal intensity, PERSI^60^). The used weighted SUVR was designed by discriminant analysis that maximally separated diagnostic groups (multiblock barycentric discriminant analysis, MUBADA^61^).

### Statistical analysis

SPSS version 24 (IBM, Armonk, NY, US), SAS (9.4) and R version 3.4.3 (RStudio)^62^ were used for statistical analysis. Group differences in the biomarker levels were assessed with Mann-Whitney test or univariate general linear models (GLM) adjusting with age and sex as covariates. Correlations between CSF biomarkers and [^18^F]flortaucipir or [^18^F]flutemetamol SUVR were examined using Pearson or Spearman tests as indicated in the Results and Figure Legends. Differences between dependent Spearman correlation coefficients were tested using methods described in Rosner et al.^63^. Diagnostic accuracies of CSF biomarkers were assessed using ROC curve analysis. AUC of two ROC curves were compared with DeLong test^64^. The [^18^F]flutemetamol SUVR cutoff >0.743 determined using mixture modeling^65^ was used to identify individuals with abnormal Aβ PET status. We also dichotomized [^18^F]flortaucipir data based on the SUVR cutoff of 1.325. P<0.05 was considered statistically significant.

## Data Availability

Anonymized data will be shared by request from any qualified investigator for the sole purpose of replicating procedures and results presented in the manuscript and as long as data transfer is in agreement with EU legislation on the general data protection regulation (GDPR).

## ACKNOWLEDGMENTS

The study was supported by the European Research Council, the Swedish Research Council, the Knut and Alice Wallenberg foundation, the Marianne and Marcus Wallenberg foundation, the Strategic Research Area MultiPark (Multidisciplinary Research in Parkinson’s disease) at Lund University, the Swedish Alzheimer Foundation, the Swedish Brain Foundation, The Parkinson foundation of Sweden, the Skåne University Hospital Foundation, and the Swedish federal government under the ALF agreement.

The funding sources had no role in the design and conduct of the study; in the collection, analysis, interpretation of the data; or in the preparation, review, or approval of the manuscript. Doses of [^18^F]flutemetamol injection were sponsored by GE Healthcare and the precursor of [^18^F]Flortaucipir was provided by AVID Radiopharmaceuticals. Euroimmun Aβ42, Aβ40 and T-tau kits were provided by Euroimmun. Analysis of p-tau217 and p-tau181 were performed at Eli Lilly and Company. We thank Dr. Wenling Zhang at Eli Lilly and Company for help with SAS statistical analysis.

## AUTHOR CONTRIBUTIONS

SJ, ES, RS, SP, NM, DCA, NKP, XC, SS, JRS, JLD, OH collected the data and reviewed the manuscript for intellectual content. SJ, DCA, JLD, and OH analyzed and interpreted the data, prepared figures and cowrote the manuscript. OH was the principal designer and coordinator of the study and overviewed collection, analysis and interpretation of the study data.

## REFERENCES

1. Scheltens, P., et al. Alzheimer’s disease. Lancet 388, 505–517 (2016).

2. Alonso, A. D., Beharry C., Corbo C. P., Cohen L. S. Molecular mechanism of prion-like tau-induced neurodegeneration. Alzheimers Dement 12, 1090–1097 (2016).

3. Hanger, D. P., Anderton B. H., Noble W. Tau phosphorylation: the therapeutic challenge for neurodegenerative disease. Trends Mol Med 15, 112–119 (2009).

4. Hernandez, F., Avila J. Tauopathies. Cell Mol Life Sci 64, 2219–2233 (2007).

5. Olsson, B., et al. CSF and blood biomarkers for the diagnosis of Alzheimer’s disease: a systematic review and meta-analysis. Lancet Neurol 15, 673–684 (2016).

6. Albert, M. S., et al. The diagnosis of mild cognitive impairment due to Alzheimer’s disease: recommendations from the National Institute on Aging-Alzheimer’s Association workgroups on diagnostic guidelines for Alzheimer’s disease. Alzheimers Dement 7, 270–279 (2011).

7. Dubois, B., et al. Advancing research diagnostic criteria for Alzheimer’s disease: the IWG-2 criteria. Lancet Neurol 13, 614–629 (2014).

8. Molinuevo, J. L., et al. Current state of Alzheimer’s fluid biomarkers. Acta Neuropathol 136, 821–853 (2018).

9. Blennow, K., Hampel H., Weiner M., Zetterberg H. Cerebrospinal fluid and plasma biomarkers in Alzheimer disease. Nat Rev Neurol 6, 131–144 (2010).

10. Buchhave, P., et al. Cerebrospinal fluid levels of beta-amyloid 1-42, but not of tau, are fully changed already 5 to 10 years before the onset of Alzheimer dementia. Arch Gen Psychiatry 69, 98–106 (2012).

11. Vos, S. J., et al. Preclinical Alzheimer’s disease and its outcome: a longitudinal cohort study. Lancet Neurol 12, 957–965 (2013).

12. Lleo, A., et al. A 2-Step Cerebrospinal Algorithm for the Selection of Frontotemporal Lobar Degeneration Subtypes. JAMA Neurol 75, 738–745 (2018).

13. Schoonenboom, N. S., et al. Cerebrospinal fluid markers for differential dementia diagnosis in a large memory clinic cohort. Neurology 78, 47–54 (2012).

14. Buerger, K., et al. CSF phosphorylated tau protein correlates with neocortical neurofibrillary pathology in Alzheimer’s disease. Brain 129, 3035–3041 (2006).

15. Tapiola, T., et al. Cerebrospinal fluid {beta}-amyloid 42 and tau proteins as biomarkers of Alzheimer-type pathologic changes in the brain. Arch Neurol 66, 382–389 (2009).

16. Brier, M. R., et al. Tau and Abeta imaging, CSF measures, and cognition in Alzheimer’s disease. Sci Transl Med 8, 338ra366 (2016).

17. Chhatwal, J. P., et al. Temporal T807 binding correlates with CSF tau and phospho-tau in normal elderly. Neurology 87, 920–926 (2016).

18. Mattsson, N., et al. (18)F-AV-1451 and CSF T-tau and P-tau as biomarkers in Alzheimer’s disease. EMBO Mol Med 9, 1212–1223 (2017).

19. Mattsson, N., et al. Comparing (18)F-AV-1451 with CSF t-tau and p-tau for diagnosis of Alzheimer disease. Neurology 90, e388–e395 (2018).

20. Gordon, B. A., et al. The relationship between cerebrospinal fluid markers of Alzheimer pathology and positron emission tomography tau imaging. Brain 139, 2249–2260 (2016).

21. Hampel, H., et al. Measurement of phosphorylated tau epitopes in the differential diagnosis of Alzheimer disease: a comparative cerebrospinal fluid study. Arch Gen Psychiatry 61, 95–102 (2004).

22. Sato, C., et al. Tau Kinetics in Neurons and the Human Central Nervous System. Neuron 97, 1284–1298 e1287 (2018).

23. Wildsmith, K. R., et al. TAU BURDEN MEASURED USING [18F]GTP1 CORRELATES WITH CSF TAU PHOSPHORYLATION AT SITES T217 AND T205 MORE CLOSELY THAN T181. Alzheimer’s & Dementia: The Journal of the Alzheimer’s Association 14, 1059–1060 (2018).

24. Ossenkoppele, R., et al. Associations between tau, Abeta, and cortical thickness with cognition in Alzheimer disease. Neurology 92, e601–e612 (2019).

25. Ossenkoppele, R., et al. Discriminative Accuracy of [18F]flortaucipir Positron Emission Tomography for Alzheimer Disease vs Other Neurodegenerative Disorders. JAMA 320, 1151–1162 (2018).

26. Janelidze, S., et al. Concordance Between Different Amyloid Immunoassays and Visual Amyloid Positron Emission Tomographic Assessment. JAMA Neurol 74, 1492–1501 (2017).

27. Xia, C. F., et al. [(18)F]T807, a novel tau positron emission tomography imaging agent for Alzheimer’s disease. Alzheimers Dement 9, 666–676 (2013).

28. Lowe, V. J., et al. An autoradiographic evaluation of AV-1451 Tau PET in dementia. Acta Neuropathol Commun 4, 58 (2016).

29. Marquie, M., et al. Validating novel tau positron emission tomography tracer [F-18]-AV-1451 (T807) on postmortem brain tissue. Ann Neurol 78, 787–800 (2015).

30. Sander, K., et al. Characterization of tau positron emission tomography tracer [(18)F]AV-1451 binding to postmortem tissue in Alzheimer’s disease, primary tauopathies, and other dementias. Alzheimers Dement 12, 1116–1124 (2016).

31. Smith, R., Wibom M., Pawlik D., Englund E., Hansson O. Correlation of In Vivo [18F]Flortaucipir With Postmortem Alzheimer Disease Tau Pathology. JAMA Neurol, (2018).

32. Zheng-Fischhofer, Q., et al. Sequential phosphorylation of Tau by glycogen synthase kinase-3beta and protein kinase A at Thr212 and Ser214 generates the Alzheimer-specific epitope of antibody AT100 and requires a paired-helical-filament-like conformation. Eur J Biochem 252, 542–552 (1998).

33. Hanger, D. P., et al. Novel phosphorylation sites in tau from Alzheimer brain support a role for casein kinase 1 in disease pathogenesis. J Biol Chem 282, 23645–23654 (2007).

34. Yoshida, H., Goedert M. Sequential phosphorylation of tau protein by cAMP-dependent protein kinase and SAPK4/p38delta or JNK2 in the presence of heparin generates the AT100 epitope. J Neurochem 99, 154–164 (2006).

35. Spillantini, M. G., Crowther R. A., Goedert M. Comparison of the neurofibrillary pathology in Alzheimer’s disease and familial presenile dementia with tangles. Acta Neuropathol 92, 42–48 (1996).

36. Braak, H., Alafuzoff I., Arzberger T., Kretzschmar H., Del Tredici K. Staging of Alzheimer disease-associated neurofibrillary pathology using paraffin sections and immunocytochemistry. Acta Neuropathol 112, 389–404 (2006).

37. Braak, H., Braak E. Neuropathological stageing of Alzheimer-related changes. Acta Neuropathol 82, 239–259 (1991).

38. Jack, C. R., Jr., et al. NIA-AA Research Framework: Toward a biological definition of Alzheimer’s disease. Alzheimers Dement 14, 535–562 (2018).

39. Blennow, K., Mattsson N., Scholl M., Hansson O., Zetterberg H. Amyloid biomarkers in Alzheimer’s disease. Trends Pharmacol Sci 36, 297–309 (2015).

40. Hansson, O., et al. The impact of preanalytical variables on measuring cerebrospinal fluid biomarkers for Alzheimer’s disease diagnosis: A review. Alzheimers Dement 14, 1313–1333 (2018).

41. Shaw, L. M., et al. Appropriate use criteria for lumbar puncture and cerebrospinal fluid testing in the diagnosis of Alzheimer’s disease. Alzheimers Dement 14, 1505–1521 (2018).

42. Petersen, R. C. Mild cognitive impairment as a diagnostic entity. J Intern Med 256, 183–194 (2004).

43. McKhann, G. M., et al. The diagnosis of dementia due to Alzheimer’s disease: recommendations from the National Institute on Aging-Alzheimer’s Association workgroups on diagnostic guidelines for Alzheimer’s disease. Alzheimers Dement 7, 263–269 (2011).

44. Gelb, D. J., Oliver E., Gilman S. Diagnostic criteria for Parkinson disease. Arch Neurol 56, 33–39 (1999).

45. Emre, M., et al. Clinical diagnostic criteria for dementia associated with Parkinson’s disease. Mov Disord 22, 1689-1707; quiz 1837 (2007).

46. Litvan, I., et al. Clinical research criteria for the diagnosis of progressive supranuclear palsy (Steele-Richardson-Olszewski syndrome): report of the NINDS-SPSP international workshop. Neurology 47, 1–9 (1996).

47. McKeith, I. G., Perry E. K., Perry R. H. Report of the second dementia with Lewy body international workshop: diagnosis and treatment. Consortium on Dementia with Lewy Bodies. Neurology 53, 902–905 (1999).

48. Lang, A. E., Riley D. E., Bergeron C. Cortical-basal ganglionic degeneration. In: Neurodegenerative Diseases (ed^(eds Calne DB). W.B. Saunders (1994).

49. Gorno-Tempini, M. L., et al. Classification of primary progressive aphasia and its variants. Neurology 76, 1006–1014 (2011).

50. Rascovsky, K., et al. Sensitivity of revised diagnostic criteria for the behavioural variant of frontotemporal dementia. Brain 134, 2456–2477 (2011).

51. Mattsson, N., et al. Predicting diagnosis and cognition with (18)F-AV-1451 tau PET and structural MRI in Alzheimer’s disease. Alzheimers Dement, (2019).

52. Honig, L. S., et al. Trial of Solanezumab for Mild Dementia Due to Alzheimer’s Disease. N Engl J Med 378, 321–330 (2018).

53. Palmqvist, S., et al. Accuracy of brain amyloid detection in clinical practice using cerebrospinal fluid beta-amyloid 42: a cross-validation study against amyloid positron emission tomography. JAMA Neurol 71, 1282–1289 (2014).

54. Hahn, A., et al. Modeling Strategies for Quantification of In Vivo (18)F-AV-1451 Binding in Patients with Tau Pathology. J Nucl Med 58, 623–631 (2017).

55. Maass, A., et al. Comparison of multiple tau-PET measures as biomarkers in aging and Alzheimer’s disease. Neuroimage 157, 448–463 (2017).

56. Cho, H., et al. In vivo cortical spreading pattern of tau and amyloid in the Alzheimer disease spectrum. Ann Neurol 80, 247–258 (2016).

57. Van Essen, D. C. A Population-Average, Landmark- and Surface-based (PALS) atlas of human cerebral cortex. Neuroimage 28, 635–662 (2005).

58. Palmqvist, S., et al. Earliest accumulation of beta-amyloid occurs within the default-mode network and concurrently affects brain connectivity. Nat Commun 8, 1214 (2017).

59. Palmqvist, S., et al. Accurate risk estimation of beta-amyloid positivity to identify prodromal Alzheimer’s disease: Cross-validation study of practical algorithms. Alzheimers Dement 15, 194–204 (2019).

60. Southekal, S., et al. Flortaucipir F 18 Quantitation Using Parametric Estimation of Reference Signal Intensity. J Nucl Med 59, 944–951 (2018).

61. Devous, M. D., Sr., et al. Test-Retest Reproducibility for the Tau PET Imaging Agent Flortaucipir F 18. J Nucl Med 59, 937–943 (2018).

62. Team, R. C. R: A Language and Environment for Statistical Computing. R Foundation for Statistical Computing, Vienna, Austria URL http://www.R-project.org/, (2014).

63. Rosner, B., Wang W., Eliassen H., Hibert E. Comparison of Dependent Pearson and Spearman Correlation Coefficients with and without Correction for Measurement Error. J Biom Biostat 6, 1–9 (2015).

64. Robin, X., et al. pROC: an open-source package for R and S+ to analyze and compare ROC curves. BMC bioinformatics 12, 77 (2011).

65. Benaglia, T., Chauveau D., Hunter D. R., Young D. S. mixtools: An R Package for Analyzing Finite Mixture Models. J Stat Softw 32, 1–29 (2009).

